# Metagenomic sequencing of municipal wastewater provides a near-complete SARS-CoV-2 genome sequence identified as the B.1.1.7 variant of concern from a Canadian municipality concurrent with an outbreak

**DOI:** 10.1101/2021.03.11.21253409

**Authors:** Chrystal Landgraff, Lu Ya Ruth Wang, Cody Buchanan, Matthew Wells, Justin Schonfeld, Kyrylo Bessonov, Jennifer Ali, Erin Robert, Celine Nadon

## Abstract

Laboratory-based wastewater surveillance for SARS-CoV-2, the causative agent of the ongoing COVID-19 pandemic, can be conducted using RT-qPCR-based screening of municipal wastewater samples. Although it provides rapid viral detection and can inform SARS-CoV-2 abundance in wastewater, this approach lacks the resolution required for viral genotyping and does not support tracking of viral genome evolution. The recent emergence of several variants of concern, a result of mutations across the genome including the accrual of important mutations within the viral spike glycoprotein, has highlighted the need for a method capable of detecting the cohort of mutations associated with these and newly emerging genotypes. Here we provide an innovative methodology for the recovery of a near-complete SARS-CoV-2 sequence from a wastewater sample collected from across Canadian municipalities including one that experienced a significant outbreak attributable to the SARS-CoV-2 B.1.1.7 variant of concern. Our results demonstrate that a combined interrogation of genome consensus-level sequences and alternative alleles enables the identification of a SARS-CoV-2 variant of concern and the detection of a new allele within a viral accessory gene that may be representative of a recently evolved B.1.1.7 sublineage.

## Introduction

In late December 2019, the World Health Organization was alerted to a cluster of atypical pneumonia cases of unknown etiology in Wuhan, Hubei Province, China [1]. Shotgun sequencing of lower respiratory tract epithelial cells obtained from two patient bronchoalveolar-lavage samples revealed a novel human beta coronavirus sharing over 85% genetic identity with a SARS-like CoV (bat-SL-CoVZC45, MG772933.1) [2]. On March 11^th^, 2020 the World Health Organization declared the rapid and global spread of the virus now designated as severe acute respiratory syndrome coronavirus 2 (SARS-CoV-2), a global pandemic. Between December 2019 and February 2021, there have been over 113 million globally confirmed cases and 2.52 million deaths (https://coronavirus.jhu.edu/map.html). At the time of preparing this manuscript, SARS-CoV-2 had led to a total of 893,518 total clinical cases and 22,304 deaths in Canada [3].

SARS-CoV-2 is the etiologic agent of COVID-19, a disease that in the majority of individuals, is associated with asymptomatic, mild or moderate symptoms that include sore throat, fatigue, runny nose, fevers or chills, cough, headache, loss of taste and smell [2],[4]. More severe cases are associated with chest pain, shortness of breath and difficulty breathing [5]. Approximately 10-15 % cases are associated with more severe symptoms of shortness of breath and difficulty breathing and as many as an estimated 4.9-6.4% of infections in Canada and the US are fatal [6]. Gastrointestinal symptoms including loss of appetite, abdominal pain, diarrhea, nausea and vomiting are reported in as many as 5-24% cases [7]–[10]. Recent studies have found that 16-19% of COVID-19 patients may only present with gastrointestinal symptoms,[8], [8], [11].

Although the exact mechanisms leading to development of gastrointestinal complications are not fully understood, studies have shown that invasive SARS-CoV-2 gains entry into the digestive tract via attachment to intestinal epithelial cells expressing the ACE-2 receptor [12]. Viable SARS-CoV-2 including RNA from SARS-CoV-2 in saliva, sputum and feces may subsequently be excreted by infected individuals into the sewage systems.

Detection of SARS-CoV-2 RNA from wastewater samples has primarily been achieved by RT-qPCR using primer/probe sets that target the genes encoding the viral nucleocapsid, spike or envelope protein[13]–[15]. Wastewater surveillance has the advantage of capturing asymptomatic cases or positive cases from individuals who do not seek testing and may detect signals of viral presence in a community at least 48 hours prior to their detection via clinical surveillance [16]. Additionally, it provides epidemiological information on a community level by averaging individual inputs into a single consensus signal that provides unique opportunities for real-time and periodical community-level epidemiological reports, assessment of interventions and prediction of possible outbreaks [17],[18].

Ongoing genome sequencing of clinical samples has provided high resolution viral sequences informing SARS-CoV-2 surveillance and genome evolution over the course of the pandemic. Global sequencing efforts have generated more than 635,000 sequence submissions to the GISAID EpiCov database and in excess of 536,000 genomes are publicly available for analysis, as of February 10^th^, 2021. This extraordinary achievement highlights the benefits of open data access and has greatly contributed to our knowledge of viral transmission dynamics, genome evolution and diversity of the SARS-CoV-2 pandemic [19]. In Canada, clinical genomic surveillance of SARS-CoV-2 has been implemented via the Canadian COVID-19 Genomics Network (CanCOGeN).

The SARS-CoV-2 genome consists of a positive sense single-stranded RNA molecule. Single-stranded RNA viruses are characterized by a high mutation rate, although the mutation rate of SARS-CoV-2 is lower than expected due to the proof-reading activities of a viral encoded proof-reading enzyme encoded by orf14a. Although most mutations do not typically have a tendency to impact disease dynamics, several mutations have been detected in the SARS-CoV-2 genome as it spread across the globe. In April 2019, one key mutation of clinical relevance was detected that resulted in a non-synonymous mutation of an aspartate to a glycine in the 614^th^ amino acid residue of the spike protein (D614G) [20]. This mutation is associated with increased transmissibility and rapidly spread across Europe, Canada, Australia and the United States. Strains carrying this mutation have mostly replaced the ancestral strain that emerged from China in December 2019. By late 2020, several variants of concern carrying numerous mutations had emerged that potentially increase viral infectivity, or reduce the response to neutralizing antibodies and multiple variants are now circulating globally.

As of March 1, 2021, three variants of concern have been identified: the B.1.1.7/501Y.V1 variant, first detected in England from a viral sequence analysis initiated in response to an unexpected rise in COVID-19 cases [21]; the B.1.351/501Y.V2 variant, first detected in South Africa [22], and P.1/501Y.V3, first detected in travelers returning to Japan from Brazil by routine airport screening of international travellers [23],[24]. The first identification of variants of concern in Canada was on December 26, 2020 from two individuals who later disclosed close contact with a traveler who had recently returned from the UK [25]. In the two months since, there have been more than 2,480 confirmed clinical cases of the B.1.1.7 variant, at least 150 confirmed cases of the B.1.351 variant and 30 cases of the P.1 variant reported in Canada [26].

In this report, we describe the first identification of a near-complete SARS-CoV-2 consensus – level genome sequence obtained from a Canadian municipal wastewater treatment plant and the detection of several mutations defining the B.1.1.7 variant of concern within the sample.

## Methods

### Wastewater Sample collection and viral concentration

A SARS-CoV-2 positive 24 hour composite influent wastewater sample from a wastewater treatment facility servicing a small Canadian municipality (pop. ∼150K) was obtained from collaborators at the University of Ottawa. The sample was collected on January 26th 2020, frozen at −80°C prior to shipment to the National Microbiology Laboratory in Winnipeg, where it was received on February 3rd 2020. A previously published allele-specific primer extension RT-qPCR assay targeting the B.1.1.7 D3L mutation within the nucleocapsid gene was used to initially screen the sample for the putative presence of the variant of concern B.1.1.7 in the wastewater sample [27].

The wastewater was processed as per Nemudryi et al. [28] with modifications. Briefly, the sample was filtered through sterile cheesecloth, and mixed vigorously prior to sequentially filtering 200 mL through a combination of 10 µm, 5 µm (Pall; Cat # 60344, 60178) and 0.45 µm membranes (VWR; Cat # CA28147-979) by vacuum filtration. The filtrate was retained and concentrated to approximately 300 µL using the Amicon Ultra-15 (10 kDa) centrifugal filter unit (Millipore; UFC9100) by ultrafiltration at 4°C and 4000 x g.

Additionally, a 30 mL aliquot of the composite influent wastewater was centrifuged at 4200 x g at 4°C for 20 minutes. The supernatant was discarded, except for approximately 500 uL used to resuspend the solids pellet.

### Nucleic Acid Extraction

Nucleic acids were extracted from the concentrated filtrate using the RNeasy Mini Kit (QIAgen; 74106) with modifications for liquid samples. Briefly, 300 µL of the concentrated filtrate was added to two volumes of RLT lysis buffer containing 2-Mercaptoethanol (1:100) and then processed according to the manufacturer’s specifications. Nucleic acids were eluted in 50 µL of nuclease-free water.

The resuspended solids pellet was added to 700-uL of RLT lysis buffer (QIAgen; 79216) containing 2-Mercaptoethanol (1:100) and bead beated using 0.7 mm ceramic garnet beads (PowerBead Tubes, QIAgen; Cat # 13123-50) on a MP Bio Fastprep-24 tissue homogenizer for 4 cycles of 20 seconds at 4.0 M/s. The sample was centrifuged at 14,000 x g for three minutes to pellet any solids, and the supernatant was retained. The supernatant was added to an equal volume of 70% ethanol and mixed thoroughly prior to extraction using the RNeasy Mini Kit (QIAgen; 74106), as per manufacturer’s specifications, beginning with the addition of the sample to the spin-column. Nucleic acids were eluted in 50 µL of nuclease-free water.

### Real-time qPCR

The presence of SARS-CoV-2 nucleic acid in the wastewater sample was confirmed by RT-qPCR using the 2019-nCoV CDC RUO Kit (IDT, 10006713) for detection of the N1 and N2 genes. The assay was performed using the TaqPath 1-Step RT-qPCR Master Mix, CG (Thermo Scientific; A15299) according to the manufacturer’s specifications on the QuantStudio 5 (Applied Biosystems) with the following parameters: UNG incubation at 25°C for 2 minutes, reverse transcription at 50°C for 15 minutes, RT inactivation and denaturation at 95°C for 2 minutes, followed by 45 cycles of 95°C for 3 seconds, 60°C for 30 seconds, and held at 4°C.

### SARS-CoV-2 cDNA and Amplicon Preparation

First strand cDNA was generated using the SuperScript™ IV First-Strand Synthesis System (Invitrogen; 18091050) according to the manufacturer’s specification with some modifications. Briefly, a total of 10 µL of RNA was combined with 1 µL each of the 10 mM dNTP mix, 50 ng/µL random hexamers and 50 µM Oligo (dT)_20_ primer, then incubated at 65°C for 5 minutes, and kept on ice until ready for use. A total of 7 µL of a mastermix comprising 4 µL of 5x Superscript IV Buffer, 1 µL of 100 mM DTT, 1 µL of Ribonuclease Inhibitor and 1 µL of Superscript IV Reverse Transcriptase was added to each sample. The samples were serially incubated at 25°C for 10 minutes, 42°C for 50 minutes, 70°C for 10 minutes, and held at 5°C. All incubations were conducted using a Mastercycler Pro thermal cycler (Eppendorf, Mississauga, ON, Canada).

The SARS-CoV-2 amplicons were generated according to the protocol described by Freed et al. [29] however the described primer set was replaced with those from the ArticV3 protocol [30], generating a series of 400-bp in lieu of 1200-bp amplicons and with additional modifications. Briefly, 100 uM stock primer pools were prepared for each set of primers (i.e., odd and even) by combining equal volumes of the appropriate primers (LabReady, 100 uM IDTE, pH 8.0; IDT) and then diluted to 10 uM prior to use. Two separate PCR reactions, one for each primer pool, were assembled comprising 5 µL of 5X Q5 Reaction Buffer, 0.5 µL of 10 mM dNTPs, 2.5 µL of cDNA, 0.25 µL of Q5 Hot Start DNA Polymerase (NEB, Massachusetts, USA; M0493S), 15.9 µL of nuclease-free water, and 1.1 µL of either 10 µM Primer Pool 1 or 2. For each pool, up to three reactions were prepared to increase yield. PCR was carried out with a denaturation stage at 98°C for 30 seconds, followed by 35 cycles of 98°C for 15 seconds and 65°C for 5 minutes, with a final hold at 4°C using the Mastercycler Pro thermal cycler (Eppendorf). The PCR products were assayed on the QIAxcel Capillary Gel Electrophoresis System (QIAgen, Mississauga, ON, CAN) with the DNA Screening Kit (QIAgen; 929004) for confirmation of successful amplification. Replicate samples were combined and concentrated using Amicon 30 kDa Ultra-0.5 Centrifugal Filter Units (Millipore, UFC503096) before quantification with the Qubit 4 fluorometer (Invitrogen) using the dsDNA High Sensitivity Kit (Invitrogen; Q32854).

### Nanopore Library Preparation and Sequencing

A total of 3.75 µL of the pooled PCR products was used as input to generate barcoded sequencing libraries using the Rapid Barcoding Kit (ONT; SQK-RBK004) according to the manufacturer’s specifications. For each sample, replicate libraries were prepared to ensure sufficient input for flow cell loading requirements. The barcoded libraries were pooled, then concentrated using an equivalent volume of AMPure XP Beads (Beckman Coulter; A63881), and resuspended in 5 µL of 10 mM Tris-HCl with 50 mM NaCl, pH 8.0 prior to completion of the library construction protocol and flow cell loading. The sequencing run was executed using MinKNOW v 3.4.1 Flongle flow cell (ONT; FLO-FLG001) on a Mk1b MinION sequencer with default settings, except live basecalling was disabled, and the .fast5 files were written to disk after every 1000 reads sequenced.

### Nanopore Sequence Data Processing

The raw reads were basecalled, trimmed and demultiplexed using Guppy v4.2.2. with the high accuracy model (dna_r9.4.1_450bps_hac.cfg). Reads from the filtrate and solids samples were first analyzed separately before being combined and analysed as a single dataset. Analysis was conducted using the Nextflow-enabled Virontus workflow v 1.1.0 [31] which automates primer trimming, read mapping, taxonomic classification of sequencing reads, consensus generation and *de novo* assembly. The SARS-CoV-2 Wuhan-Hu-1 (GenBank accession number MN908947.3) reference sequence and primer .bed file used for these analyses were provided by the artic-network primer schemes for SARS-CoV-2 available on GitHub [32]. The clade and lineage assignments for the wastewater consensus sequence, herein designated as 21-WW-BAR-01, were determined using the NextClade [33] and Pangolin [34] web applications respectively. Summary quality statistics were calculated using the basecalled .fastq file output by Guppy using NanoPlot [35] with default settings.

### Data retrieval, Sequence Alignment and Phylogenetic Analysis

A multiple sequence alignment (MSA) was generated comprising all high-quality and deduplicated (i.e., <5% ambiguous base calls) SARS-CoV-2 sequences (n=459,421) retrieved from the GISAID EpiCoV database on February 10th, 2021. Sequences representing the human clinical isolates from Canada (n=9,189) were retrieved. To reduce the computational resources required for phylogenetic analysis, the Canadian sequences were subsampled by randomly selecting two sequences per Pangolin lineage from each Canadian province. In cases where there was a single representative sequence or no province or territory was provided, the sequence was retained. After subsampling, only four sequences belonging to the B.1.1.7 lineage were present in the Canadian dataset. To increase confidence in the placement of the wastewater consensus sequence in the phylogeny, additional B.1.1.7 sequences were retrieved from GISAID’s MSA by randomly selecting one representative sequence from each country excluding Canada. The subsampled Canadian (n=536) and B.1.1.7 (n=49) sequences, wastewater consensus sequence and SARS-CoV-2 Wuhan-Hu-1 (MN908947.3) reference sequence were re-aligned using MAFFT v7.475 with the -add command using default settings [36], [37]. The resultant MSA was used to generate a maximum likelihood tree with the IQ-TREE COVID-19 release v 2.1.2 [38] using default parameters, automatic model selection, ultrafast bootstrapping approximation with 1000 replicates, and the SARS-CoV-2 Wuhan-Hu-1 original sequence as the outgroup. The phylogenetic trees were visualised using the ggtree package [39] with branches coloured based on clade designation as determined by Nextclade.

### Variant analyses

Variant analysis was conducted on the trimmed .bam files outputted by the Virontus pipeline using SAMtools v 1.7 [40] and iVar v 1.3 [41] as described in their documentation files, except the minimum quality score used to determine whether or not to count a base (-q) was set to 10, the minimum frequency threshold (-t) was set to 0.01, and the minimum read depth to call variants (-m) was set to 10. The fasta and GFF3 files corresponding to SARS-CoV-2 Wuhan-Hu-1 (Genbank accession number MN908947.3) were used as references. Variants identified by iVar were screened for mutations associated with the VOCs. Additionally, all variants positions were interrogated for possible assignment to the B.1.1.7 lineage, excluding those representing frameshift mutations (i.e., indels of n length not divisible by three).

## RESULTS AND DISCUSSION

### Wastewater collection from a municipality experiencing an outbreak of B.1.1.7

At the time the wastewater sample was collected, the community was experiencing an outbreak in a personal care home that resulted in SARS-CoV-2 being diagnosed in all but one resident [42]. Between Jan 8 and Feb 18 when the outbreak was declared over, at least 244 individuals had contracted the virus, including 129 residents, 106 staff members, 69 residents and 1 care worker had succumbed to COVID-19 [42]. Genome sequencing of oral nasopharyngeal swabs revealed that the outbreak was associated with a variant of concern, B.1.1.7/N501Y.V1 [27], [43]. This variant is distinguished by a number of mutations from the ancestral Wuhan Hu-1 “wild-type” strain and is associated with increased transmissibility [43], increased disease severity [44] and potentially increased fatality [44]. These characteristics are thought to be attributed to clinically significant mutations within the spike protein: N501Y a key residue in the receptor binding domain (RBD) that is thought to increase ACE2 receptor affinity [45] and P681H, a residue in the furin cleavage site between S1 and S2 subunits in spike [46]. The S1/S2 furin cleavage site is unique to SARS-CoV-2 compared to other members of the betacoronavirus family and has been demonstrated to promote fusion between SARS-CoV-2 and host cell membranes [47]. Mutations in this region may promote viral entry into epithelial cells [48], increase transmissibility [49] or modulate the host immune response [46], [50].

### Determination of Ct values to inform the recovery of SARS-CoV-2 from the wastewater sample

Previous wastewater sequencing experiments within our laboratory suggested an association between Ct values and breadth of genome coverage achievable with SARS-CoV-2 sequencing; we have found that wastewater samples with Ct values greater than 36-37 have low probability of providing near-complete genome coverage upon sequencing (data not shown). Additionally, a previously published study observed an inverse sigmoidal correlation between N1 and N2 Ct values and the percentage of the genome recovered using Nanopore sequencing [51]. To increase the likelihood of obtaining a near-complete SARS-CoV-2 genome, we therefore proceeded to sequence both the solids and the fltrate fractions of the wastewater sample.

Detection of the N1 and N2 genes by RT-qPCR resulted in Ct values of 37.6 and 36.1 for the filtrate sample, and 34.5 and 34.6 for the solids sample. This corresponds to an estimated 2-20 RNA copies/µl in the wastewater sample based on the standard curve determined in-house using the SARS-CoV-2 control plasmid (IDT).

### Phylogenetic analysis of the wastewater consensus sequence

Wastewater comprises inputs from many individuals, and therefore, the SARS-CoV-2 consensus sequence generated from this sample is presumably representative of either a dominant strain, or a composite of multiple strains circulating at the time of sampling.

When analyzed either as contextualized amongst a representative set of Canadian and B.1.1.7 sequences using maximum likelihood and the GTR+F+R2 evolutionary model, or when placed into a representative global phylogeny using Nextclade’s Clade Assignment tool, both approaches placed our wastewater consensus sequence in the 20I/501Y.V1 clade. This was evidence for the presence of this VOC in the sample, though its placement within the clade differed between the analyses. In the maximum likelihood tree (Figure 1), the consensus sequence was placed earlier in the clade, whereas within the topology of the Nextclade phylogeny (data not shown), it was placed more centrally. This disparity may likely be attributed to several factors including the use of different datasets for the analyses, differences in algorithms implemented in each of the software used to reconstruct the phylogenies, as well as interpretation of the consensus sequence itself, which likely comprises signal from multiple SARS-CoV-2 lineages.

**Figure 1.**
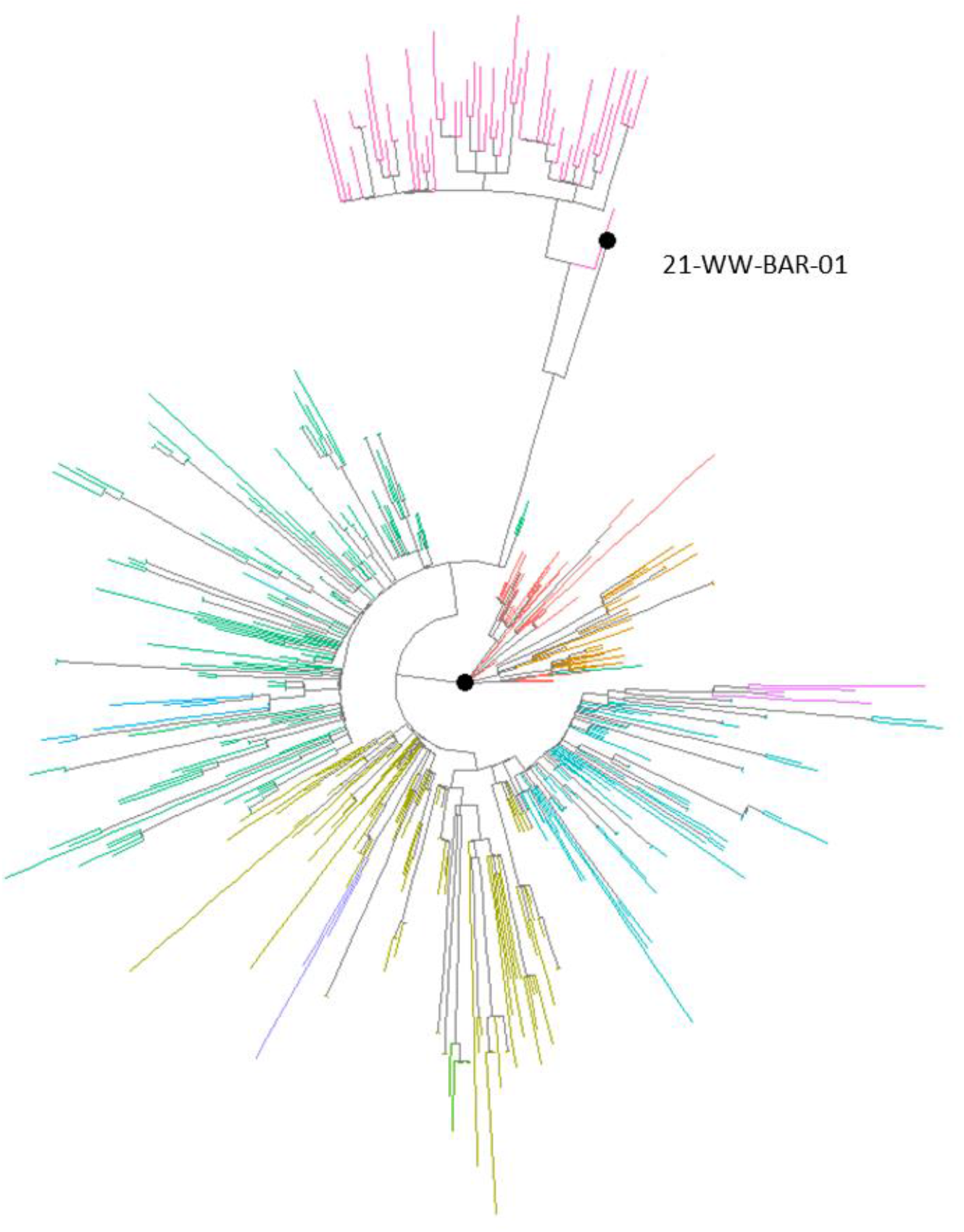
Maximum-Likelihood Tree of 536 Canadian sequences and 49 B.1.1.7 sequences sampled from the GISAID EpiCov database. Tree is rooted to the Wuhan Hu-1 reference genome (GenBank accession # MB908947.3). Branches are coloured according to assigned Nextclade lineages. The position of the wastewater consensus sequence, 21-WW-BAR-01, is indicated on the tree and represented by a black node.

### Generation of a near-complete genome of SARS-CoV-2 VOC B.1.1.7 from wastewater

The use of wastewater genomics to help elucidate the introductory events of different viral lineages or variants into a community requires a complete or near-complete consensus sequence of the viral genome for comparison to clinical sequences. Here, we applied a previously-published tiled amplicon sequencing method supported with ONT’s MinION MK1B sequencing platform and Flongle flow cell (ref needed for tiled amplicon). A total of 285,636 reads were generated over a 24 hour period with an average length and quality score of 594 bases and 10, respectively. Initially, the filtrate and solids samples were analysed separately. However, after data processing, it was determined that the majority of the reads mapping to SARS-CoV-2 were generated from the solids sample. Furthermore, after demultiplexing, the barcode for 29.7% of the reads could not be accurately determined, and therefore, these reads could not be attributed to either sample. Thus, the decision was made to process the entirety of the sequencing run as a single dataset to maximize the available data. A total of 84,078 (29%) reads were mapped to the SARS-CoV-2 Wuhan-Hu-1 reference sequence resulting in an average depth of coverage of 672 X for the 93% of the genome with > 3x coverage. (Figure 2). The following regions could not be assessed for variation due to insufficient coverage: 1-41, 3522-3799, 3803, 4041-4305, 12247-12428, 13375-13610, 13975-14218, 21163-21264, 21271, 21319, 21345 21400-21402, 21409, 21438-21656, 21660, 23204-23452, 29857-29903. The recovery of near complete SARS-CoV-2 genomes from wastewater can be highly sample dependent as demonstrated by Izquierdo-Lara et al [51], where over half of the samples sequenced by Nanopore achieved less than 50% breadth of coverage. Even with these regions of insufficient coverage, the approach described in this manuscript enabled the assembly of a near-complete SARS-CoV-2 wastewater consensus sequence surpassing 93% breadth of genome coverage enabling its subsequent genomic placement within the B.1.1.7 variant of concern lineage.

**Figure 2:**
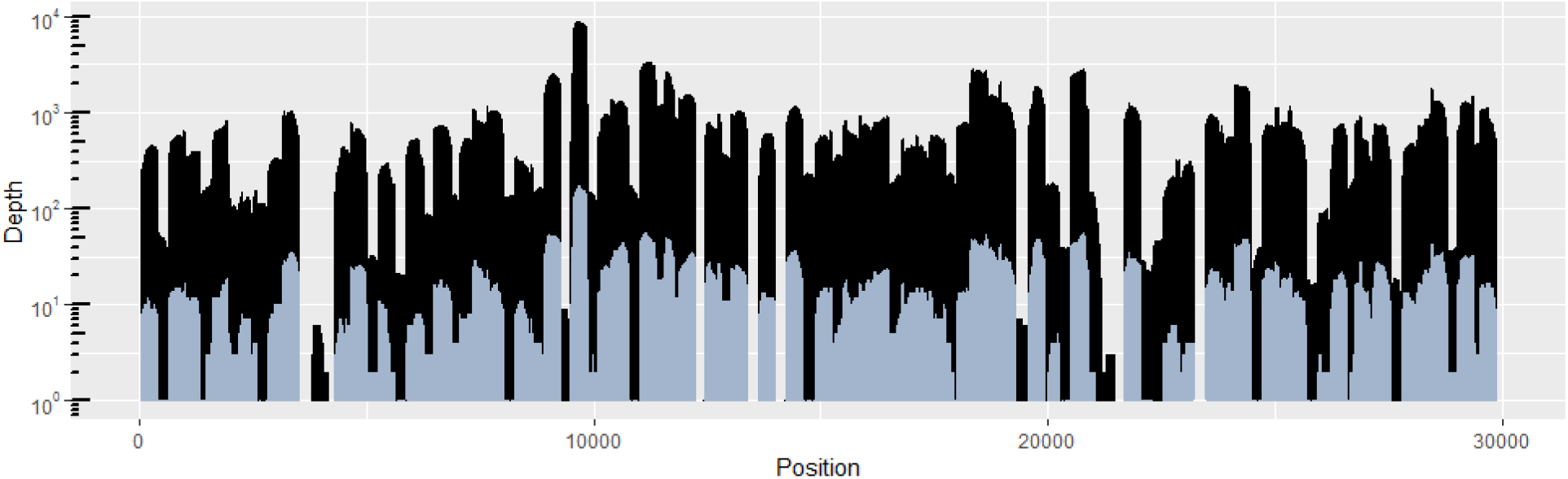
The SARS-CoV-2 genome coverage plots after 15 minutes of nanopore sequencing (blue) and after completion of the sequencing run (black). The y-axis, representing the depth of coverage, is expressed in logarithmic scale.

### Consensus level analysis reveals the presence of key B.1.1.7 mutations

The wastewater consensus sequence was assigned to clade 20I/501Y.V1, using the Nextclade Clade Assignment web tool which represents a group of isolates derived from ancestral clade 20B that carry the key B.1.1.7 VOC mutations affecting amino acid residues in the spike gene (N501Y, 570S, P681H) and Orf8 (Q27*). Nextclade analysis further revealed 24 mutations representing divergence from the ancestral sequence, Wuhan-Hu-1 (Table 1). Several of these mutations are signature SNVs of B.1.1.7, which is delineated by 14 non-synonymous mutations, 6 synonymous mutations and 3 deletions. Ten of the B.1.1.7 signature mutations were identified in the consensus sequence including a deletion of 6 nucleotides in the spike gene (21765-21770 nt) leading to the loss of a histamine and a valine residue (del 69-70) in the spike protein and non-synonymous amino acid altering mutations in the spike protein (N501Y, S982A, D618H). For a detailed list of mutations observed in the consensus sequence see Table 1, Figure 5 and Figure 6. The N501Y mutations in the spike protein is of high biological concern due to studies demonstrating that the N501Y mutation increases viral affinity for the host ACE2 receptor [45] and may impact vaccine efficacy by reducing the effect of neutralizing antibodies [52]. The orf8 gene encodes an accessory protein proposed to act as a highly immunogenic immunoglobulin-like protein that interferes with host immune responses [53], however, the effect of mutations within the orf8 gene have not been as well characterized Three Orf8 mutations associated with the B.1.1.7 lineage were identified in the wastewater consensus sequence; Q27* and R52I and Y73C. The Q27 stop-gain mutation leads to a truncated protein which has been suggested either leads to a loss of functional activity or promotes the accrual of downstream mutations [54]. The R52I mutation is localized at a protein dimerization interface that together with the Y73C residue enable formation of Orf8 protein complexes [54]. These complexes may enable viral escape from the host immune responses [53], [55].

**Table 1:** Identification of non-synomymous signature mutations of the B.1.1.7 variant of concern and observed variant frequencies in wastewater at the consensus and subconsensus levels.

| Nucleotide position<sup>a</sup> | Amino acid residue | ORF | Variant Frequency<sup>b</sup> |
| --- | --- | --- | --- |
| C3267T | T1001I | Orf1ab | 0.013 |
| C5388A | A1708D | Orf1ab | 0.017 |
| T6954C | I2230T | Orf1ab | n/d |
| del:11288:9 | SGF 3675-3677 | Orf1ab | 0.22 |
| del:21765:770 | HV 69/70 del | spike | 0.26 |
| del:21991:3 | Y144 del | spike | 0.49 |
| **A23063T** | **N501Y** | **spike** | **0.59** |
| C23271A | A570D | spike | n/c |
| C23604A | P681H | spike | 0.43 |
| C23709T | T716I | spike | 0.42 |
| **T24506G** | **S982A** | **spike** | **0.86** |
| **G24914C** | **D1118H** | **spike** | **0.97** |
| **C27972T** | **Q27\*** | **Orf8** | **0.98** |
| **G28048T** | **R52I** | **Orf8** | **0.67** |
| **A28111G** | **Y73C** | **Orf8** | **0.83** |
| **28280 GAT>CTA** | **D3L** | **N** | **0.75** |
| **C28977T** | **S235F** | **N** | **1** |

<sup>a</sup> Relative to the SARS-CoV-2 Wuhan-Hu-1 reference sequence (GenBank accession # MN908947.3)
<sup>b</sup> Using iVar v.1.3 with minimum quality score of 10, the minimum frequency threshold of 0.01, and minimum read depth to call variants of 10. **Bolded**: consensus level (frequency > 0.50) variants.
n/d Not detected
n/c No coverage

### Identification of B.1.1.7 signature mutations by interrogation of subconsensus alleles

In addition to the identification of SNVs distinguishing B.1.1.7 at the consensus level, signature variants of this lineage were identified by analysis of subconsensus variant calls. Table 1 shows the frequency of detected variants associated with lineage B.1.1.7 sequences. The variant frequency calls for each signature SNV ranged from 59-100% of total reads with read depths of 18-542 X for variants included in the consensus sequence. For signature SNVs identified at a subconsensus threshold of less than 50% of the total SNV calls, the B.1.1.7-distinguishing SNVs represented 1 to 43% of the total SNV calls with read depths of 10 to 43 reads per variant.

Several important mutations within the spike gene were identified as alternative alleles in the subconsensus sequences including a 3 nucleotide deletion at position 21991-21993 resulting in removal of a tyrosine residue at the 144^th^ residue to the spike protein (Y144), a C23604A resulting in replacement of proline with a histidine residue (P681H) and a C23709 mutation replacing a threonine with an isoleucine (T176I). These mutations are each of potential biological relevance. Together with N501Y and 69-70del, P681H is one of three mutations in the spike gene that are hypothesized to have the greatest biological impact [56]. P681H is located near the important S1/S2 furin cleavage site required for viral fusion with the host cell membrane and mutation in this loci are hypothesized to impact viral transmissibility and enhance systemic infection [48],[49]. This mutation has been observed with increasing frequency in the GISAID database in global sequences outside of the B.1.1.7 lineages suggesting occurrence via convergent evolution [46].

### Identification of additional important mutations including a recently emerged mutation representing a sub-lineage of B.1.1.7

Analysis of the consensus level variants identified using NextClade led to the identification of three of 4four specific SNVs associated with the ancestral 20B lineage. Due to high pairwise allelic co-occurrence, C241T (5′UTR), C3037T (nsp3 F924F), C14408T (nsp12 P4715L), and A23403G (spike protein D614G) have mostly replaced the ancestral alleles and are now dominant within the globally circulating viral lineages [19]. The A23403G (D614G) mutation in the spike gene was not identified at the consensus level due to a lack of sequencing coverage at this locus.

Intriguingly, one SNV was detected in the consensus sequence that demonstrated a strong association with the B.1.1.7 lineage that has not been previously mentioned in the literature. This mutation, A28095T introduces a stop codon into a lysine at residue 68 of Orf8. Further interrogation using the Nextstrain genotyping tool and COVID-CG [57] tool, which both mine a representative subset of data from the GISAID database, revealed that this SNV appears to have recently emerged (Figure 3, Table 2). A thorough analysis of this SNV using COVID-CG indicated that this mutation was first detected in a sequence from South America collected in May 2020 and increased in frequency worldwide between October and December 2020. As further evidence supporting the detection of the B.1.1.7 lineage from the wastewater sample, this SNV has high co-occurrence in GISAID with at least 3 other mutations in Orf8 (Q27*, R52I and Y73C) in all global regions (Figure 4). Orf8 is a small accessory protein with a length of 121 amino acids which has been proposed to enable evasion of host immune responses [53], [58]. The impact of the K68 stop-gain mutation has not yet been determined.

**Figure 3:**
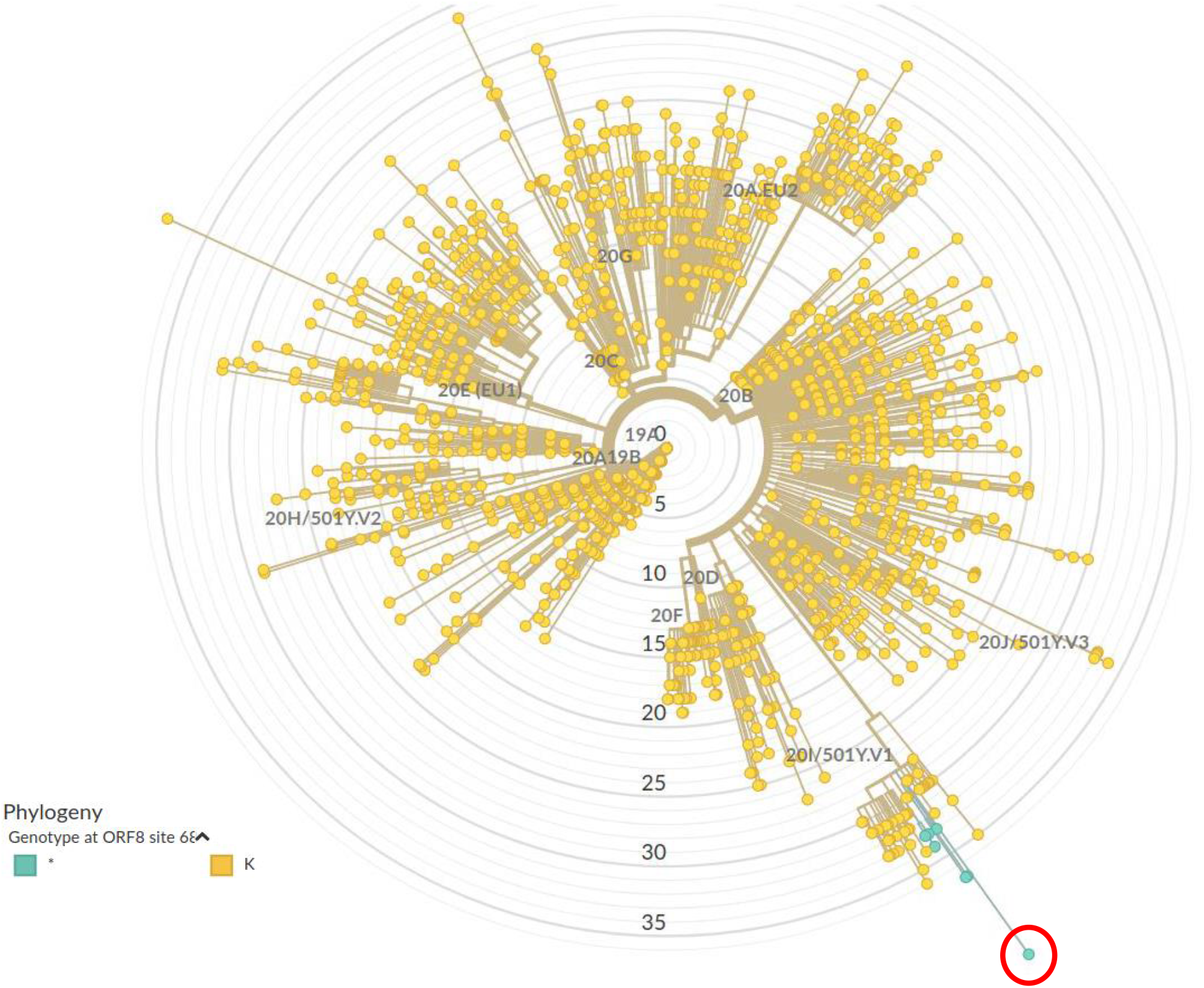
Detection of A28095T SNV in the wastewater consensus sequence that is unique to a recently emerged sublineage of B.1.1.7 sequences. The node corresponding to the wastewater sequences is circled in red.

**Table 2:**
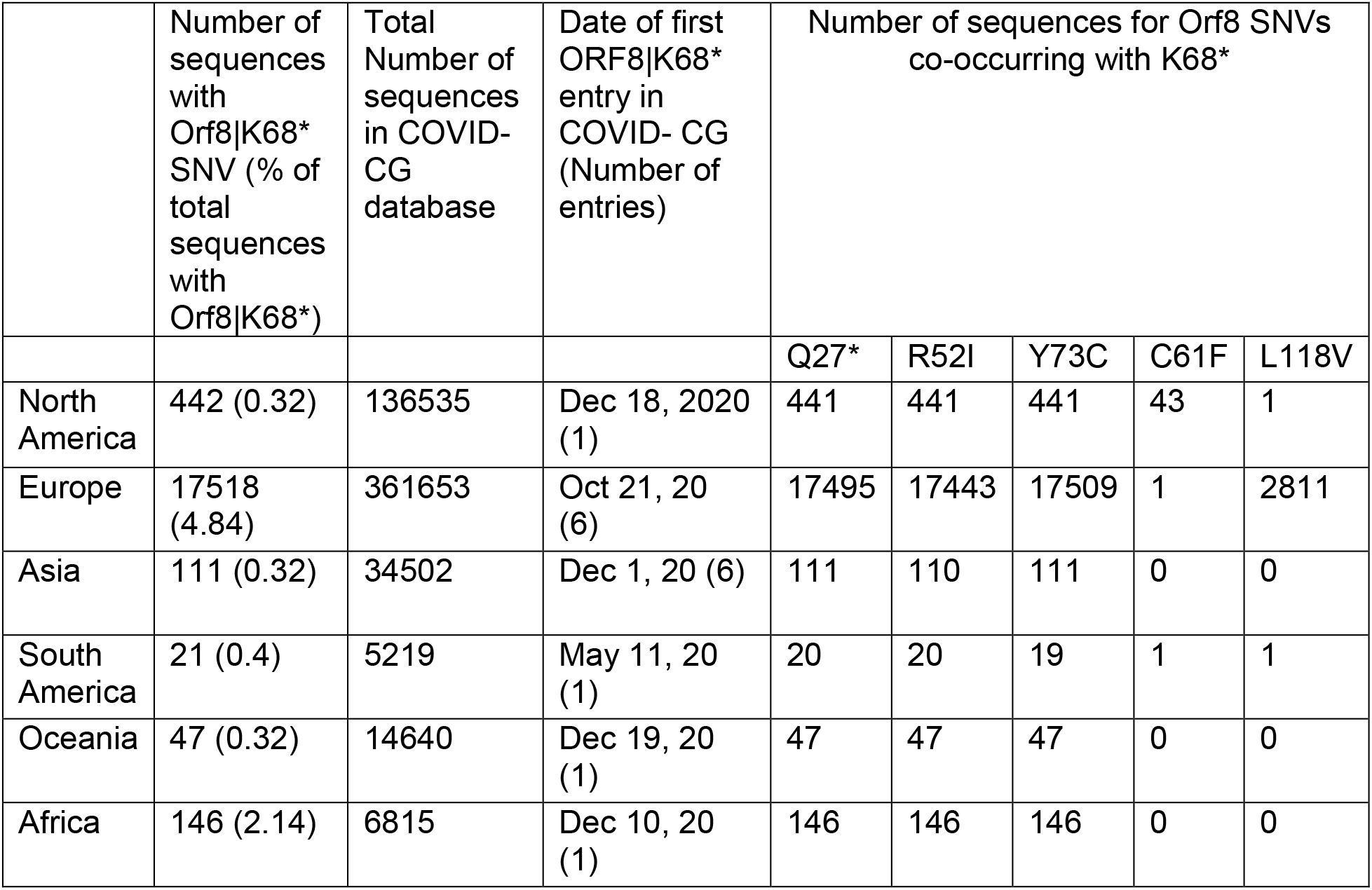
Frequency of SNVs co-occurring with Orf8 K68* in sequences deposited to GISAID identified using COVID-CG.

|  | Number of sequences with Orf8\|K68* SNV (% of total sequences with Orf8\|K68*) | Total Number of sequences in COVID-CG database | Date of first ORF8\|K68* entry in COVID-CG (Number of entries) | Number of sequences for Orf8 SNVs co-occurring with K68* |  |  |  |  |
| --- | --- | --- | --- | --- | --- | --- | --- | --- |
|  |  |  |  | Q27* | R52I | Y73C | C61F | L118V |
| North America | 442 (0.32) | 136535 | Dec 18, 2020 (1) | 441 | 441 | 441 | 43 | 1 |
| Europe | 17518 (4.84) | 361653 | Oct 21, 20 (6) | 17495 | 17443 | 17509 | 1 | 2811 |
| Asia | 111 (0.32) | 34502 | Dec 1, 20 (6) | 111 | 110 | 111 | 0 | 0 |
| South America | 21 (0.4) | 5219 | May 11, 20 (1) | 20 | 20 | 19 | 1 | 1 |
| Oceania | 47 (0.32) | 14640 | Dec 19, 20 (1) | 47 | 47 | 47 | 0 | 0 |
| Africa | 146 (2.14) | 6815 | Dec 10, 20 (1) | 146 | 146 | 146 | 0 | 0 |

**Figure 4:**
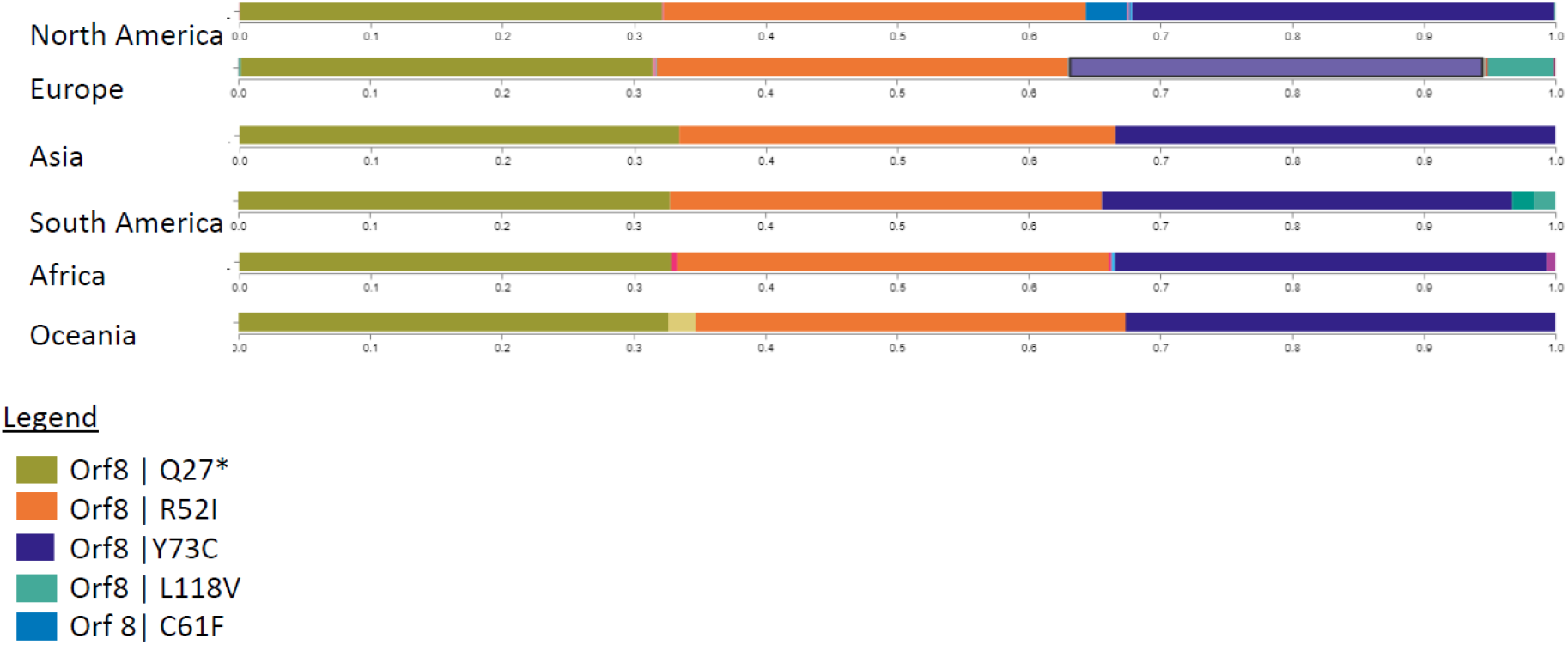
Frequency of SNVs co-occurring with Orf8 K68* according to geographical region. The plot shows the SNVs that are variable from the reference sequence WIV04 (Genbank accession #MN996528.1) and the observed frequency of each SNV normalized to the frequency observed with the K68 stop-gain mutation in the Orf8 gene arising from an A28095T mutation. The plot is modified from the COVD-CG website.

**Figure 5:**
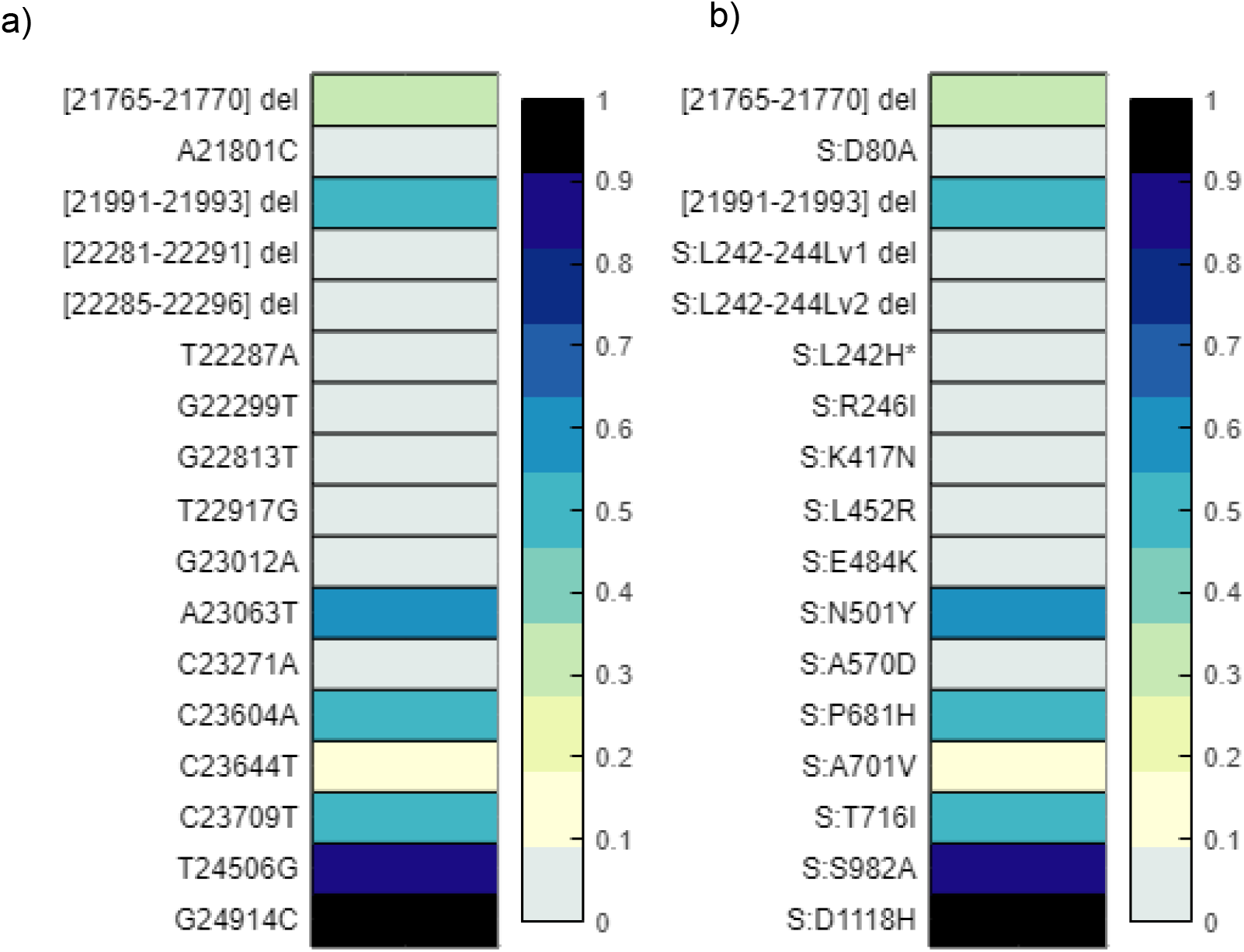
Discretized frequency of spike gene and protein mutations associated with variants of concern detected in the wastewater samples. Alleles and the frequency of each was detected using iVar. Frequencies of each signature mutation are rounded up to the nearest tenth, with values less than 0.01 indicated as a 0 or non-detects as they were determined to be below an acceptable threshold of identification. a) depicts the nucleotide changes found in the Spike gene b) is a duplicate of a) only depicting the amino acid changes resulting from each genome mutation.

**Figure 6:**
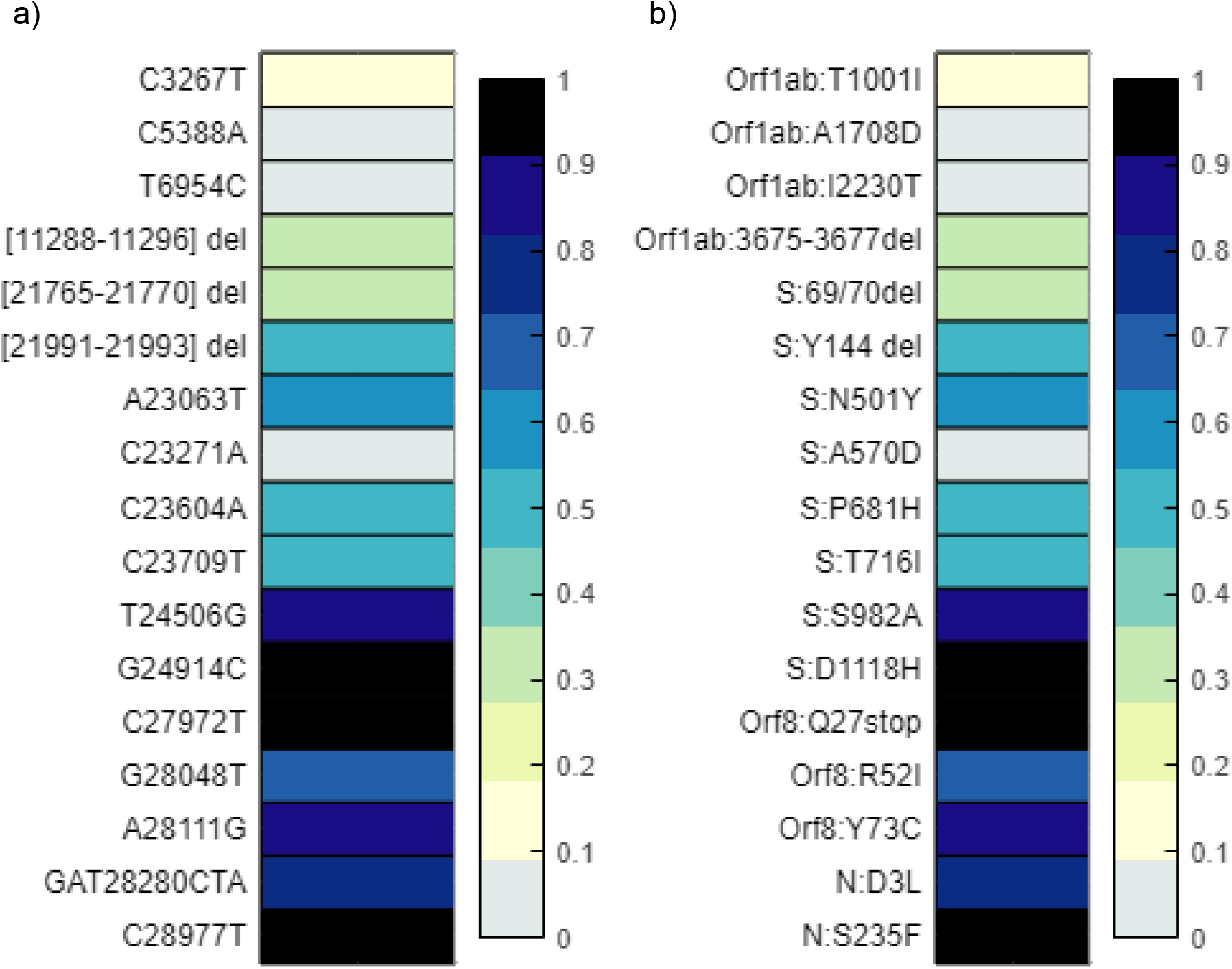
Discretized frequency of signature gene and protein mutations of lineage B.1.1.7 detected in the wastewater samples. Alleles and the frequency of each was detected using iVar. Frequencies of each signature mutation are rounded up to the nearest tenth, with values less than 0.01 indicated as a 0 or non-detects as they were determined to be below an acceptable threshold of identification. a) depicts the nucleotide changes associated with B.1.1.7 b) is a duplicate of a) only depicting the amino acid changes resulting from each genome mutation.

### Near real-time data analysis enables rapid confirmation for the presence of the B.1.1.7 VOC

To facilitate rapid confirmation of the presence of the B.1.1.7 variant, we analysed the first 5000 reads generated after only 15 minutes of sequencing. In total, 1,823 reads were mapped to the SARS-CoV-2 reference, resulting in 14 X average coverage across 70% of the genome (Figure 2). In total, 13 SNVs and three deletions distinguishing the B.1.1.7 VOC were observed with frequencies and coverage ranging from 7-100% and 1-47 X, respectively (data not shown).

## Conclusions

In this study we provide a methodology enabling sufficient viral recovery from wastewater to achieve a near-complete SARS-CoV-2 genome using Oxford Nanopore technology. Moreover, we have described an analytical approach and interpretation of wastewater data that incorporates consensus and subconsensus variant calls to detect the presence of known lineages of SARS-CoV-2 and to identify new or ng SNVs defining a specific lineage. One of the unique features of ONT sequencing is the ability to analyse data in near real-time. Using this approach, we were able to detect the presence of the B.1.1.7 variant only 15 minutes into the sequencing run. It is interesting to note that although the completed sequencing run resulted in superior genomic coverage and depth, a majority of SARS-CoV-2 genome was obtained relatively quickly. Analysis of reads from early stages of the sequencing run can be a useful mechanism, not only for gauging the success of the run, but also to provide timely results in urgent situations.

To the best of our knowledge, this study is the first to perform metagenomic sequencing of a Canadian wastewater sample for generation of a near-complete SARS-CoV-2 consensus sequence (93%). Multifaceted analysis of ONT sequencing data enabled the identification of the SARS-CoV-2 B.1.1.7 lineage in wastewater concurrent with an outbreak of the variant in a Canadian municipality. The detection of several signature mutations at the consensus and subconsensus levels supports the role of wastewater-based genomic surveillance to monitor and track the spread of SARS-CoV-2 and its variants of concern within Canada.

## Data Availability

This manuscript acknowledges the sampling and use of data obtained from the GISIAD database. The Oxford Nanopore sequencing reads generated in this study were deposited into NCBI BioProject PRJNA708265 and BioSample accession SAMN18228019.

https://www.gisaid.org/

## Date Availability

The Oxford Nanopore sequencing reads were deposited into NCBI BioProject PRJNA708265 and BioSample accession SAMN18228019

## Author Contributions

C.L, L.Y.R.W, C.B conceptualized the study. L.Y.R.W, CB J.A, and E.R conducted the methodology, M.W, J.S, L.Y.R.W, C.B, K.B and C.L. conducted bioinformatic analysis, developed analytical and visualization tools, C.L, R.W, C.B, M.W, J.S, and K.B drafted the manuscript, and C.L, L.Y.R.W and C.N compiled the final manuscript.

## Acknowledgements

The authors would like to extend their thanks to Robert Delatolla (University of Ottawa, Ottawa, ON Canada),Tyson Graber (Children’s Hospital of Eastern Ontario, Ottawa, ON Canada) and Lawrence Goodridge (University of Guelph, Guelph, ON Canada) for arranging collection and providing access to the wastewater sample as well as the greater scientific community who have been submitting data to the GISAID database.

## Competing Interests

The authors declare no competing interests.

## Notes

### Competing Interest Statement

The authors have declared no competing interest.

### Funding Statement

This study was conducted using funding provided by the Public Health Agency of Canada ( internal funding only)

### Author Declarations

This study did not require ethics approval

